# A Cross-sectional Study on Perceived Workplace Health Support and Health-Related Quality of Life

**DOI:** 10.1101/2021.09.01.21262994

**Authors:** Kazushirou Kurogi, Kazunori Ikegami, Hisashi Eguchi, Mayumi Tsuji, Seiichiro Tateishi, Tomohisa Nagata, Shinya Matsuda, Yoshihisa Fujino, Akira Ogami, the CORoNaWork Project

**Author notes:** Corresponding Author: Akira Ogami, MD, Phd, Prof., 1-1, Iseigaoka, Yahatanishi-ku, Kitakyushu, Fukuoka 807-8555, Japan. **Author contributions** KK wrote the manuscript and analyzed the data. KI reviewed the manuscript, created the questionnaire, analyzed the data, and provided advice on interpretation. AO reviewed the manuscript, analyzed the data, and provided advice on interpretation, HE,□MT,□ST,□TN,□SM reviewed the manuscript. YF reviewed the manuscript and contributed to the overall survey planning, creating the questionnaire, and securing funding for research.

## Abstract

**Objective:** Many companies in Japan have been increasingly interested in “health and productivity management (H&PM).” In terms of H&PM, we supposed that companies can enhance their employees’ perceived workplace health support (PWHS) by providing support for workers’ lively working and healthy living. This could then improve health-related QOL (HRQOL) by increasing PWHS. This study explored the relationship between PWHS and health-related quality of life (HRQOL).

**Methods:** During the COVID-19 pandemic in December 2020, we conducted an Internet-based nationwide health survey of Japanese workers (CORoNaWork study). A database of 27,036 participants was created. The question regarding the intensity of PWHS was measured using a four-point Likert scale. We used a linear mixed model (LMM) to analyze the relationship between the intensity of PWHS and the four domains of CDC HRQOL-4 (self-rated health, number of poor physical health days, number of poor mental-health days, and activity limitation days during the past 30 days).

**Results:** In the sex- and age-adjusted and multivariate models, the intensity of PWHS had a main effect on self-rated health and the three domains of unhealthy days (physical, mental, activity limitation). There was also a trend toward worse HRQOL scores as the PWHS decreased.

**Conclusions:** This study aimed to document the relationship between PWHS and HRQOL. We found that the higher the PWHS of Japanese workers, the higher their self-rated health and the lower their unhealthy days. Companies need to assess workers’ PWHS and HRQOL and promote H&PM. H&PM is also necessary to maintain and promote the health of workers during the COVID-19 pandemic.

## Introduction

Japan is facing a decline in workers, an aging population, and reduced productivity. ^1^ As one solution to these problems, companies should be proactively involved in maintaining and promoting the health of their employees. In recent years, many companies have become more interested in “health and productivity management (H&PM),” which is an employee health management approach from a corporate management perspective, and have strategically promoted it.^2,3^

To promote H&PM, companies should both reinforce workplace health support and consider how workers perceive their efforts. The concept of perceived organizational support (POS) is known as workers’ expression of evaluations and perceptions of the organization. POS was proposed by Eisenberger et al. in 1986 and is defined as “global beliefs concerning the extent to which the organization values their contributions and cares about their well-being”.^4, 5^ In terms of H&PM, we supposed that companies can enhance their employees’ perceived workplace health support (PWHS) by providing support for workers’ lively working and healthy living. Prior studies have reported that PWHS can be measured by employees’ POS for ensuring healthy living and engagement in physical activity.^6^

The Coronavirus disease 2019 (COVID-19) has a greater impact on physical and mental health due to changes in work practices, including infection prevention, than acute respiratory symptoms due to infection.^7,8^ During the COVID-19 pandemic, it has been reported that a high proportion of people have experienced mental and emotional deterioration and that these adverse psychological effects are associated with physical and social inactivity, poor sleep quality, unhealthy eating habits.^9,10^ These effects will be reflected in Quality of life (QOL), which usually includes subjective evaluations of positive and negative aspects of life.^11^ The current COVID-19 pandemic may negatively impact workers’ QOL in health-related domain and reduce their productivity at work. Maintaining or improving health-related QOL (HRQOL) through workplace health support may be important.

Few studies have focused on PWHS, and none have examined the relationship between PWHS and HRQOL. We focused on these to clarify the relationship between PWHS and workers’ health status by using a large-scale internet survey of workers conducted during the COVID-19 pandemic (December 2020).

## Methods

### Study Design and Setting

A prospective cohort study was conducted by the research group from the University of Occupational and Environmental Health, Japan, called Collaborative Online Research on Novel-coronavirus and Work study (CORoNaWork study). This study was a self-administrated questionnaire survey conducted by a Japanese online survey company (Cross Marketing Inc. Tokyo), and a baseline survey was conducted from December 22 to 25, 2020. This study design is a cross-sectional study using a part of a baseline survey of the CORoNaWork study. Fujino et al. introduced the details of this study protocol. ^12^ This study was approved by the Ethics Committee of the University of Occupational and Environmental Health, Japan.

### Participants

During the baseline survey, participants were between 20 and 65 years of age and working. A total of 33,087 participants, stratified by cluster sampling by gender, age, region, and occupation, participated in the CORoNaWork study. A database of 27,036 participants was created by excluding 6,051 with invalid responses (Fig 1).

**Fig. 1.**
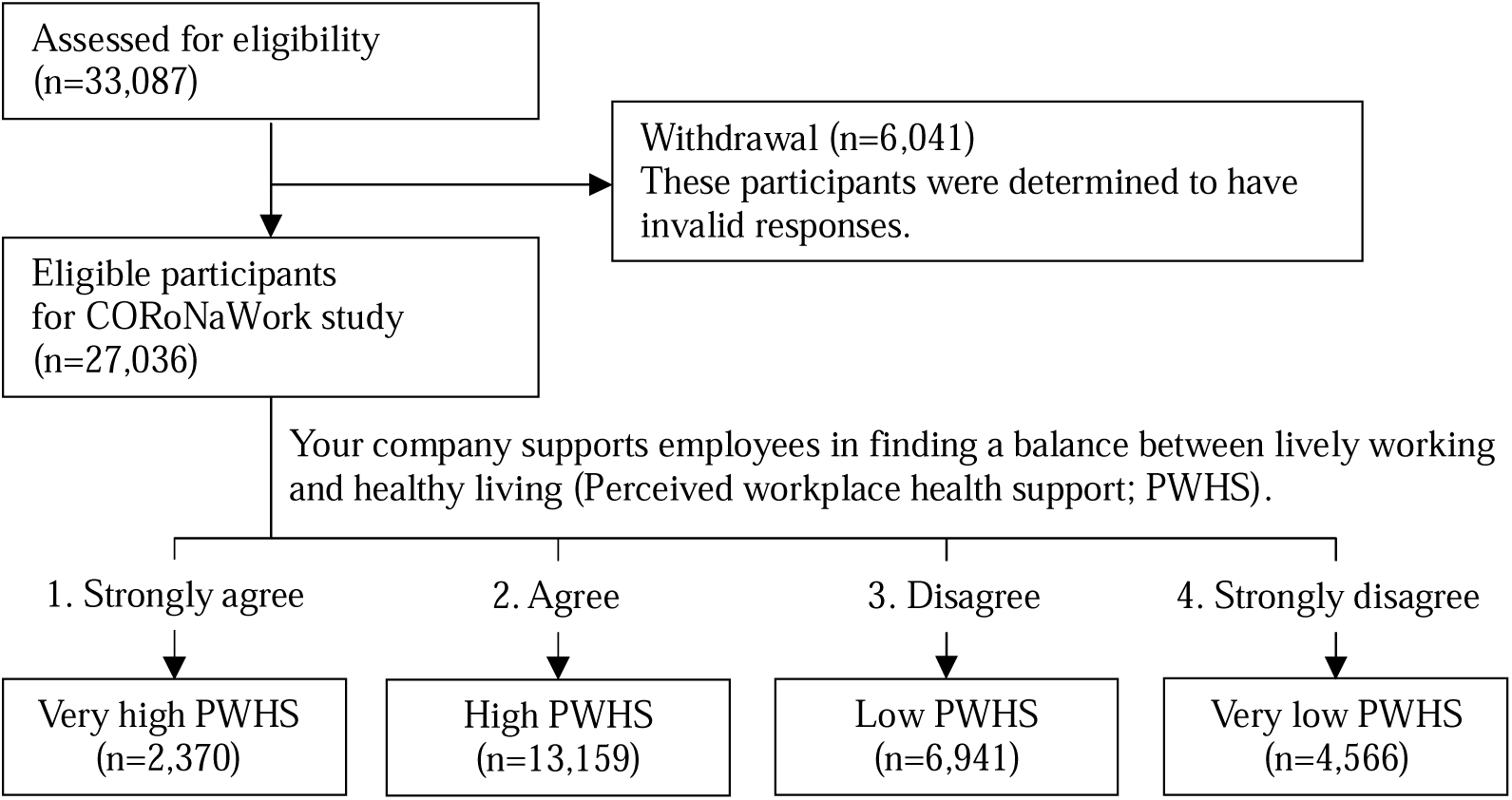
Flow chart of this study population selection.

### Questionnaires

The questionnaire items used in this study are described in detail by Fujino et al.^12^ We used data on sex, age, educational background, presence of illnesses that require hospital treatment, job type, company size where participants work, working hours per day, and work-related data.

One measure of HRQOL is the HRQOL-4 developed by the Centers for Disease Control and Prevention (CDC) (hereafter CDC HRQOL-4).^13,14^ It fetches a self-rated health status by asking about the following four domains: (a) self-rated health (5-Point Likert Scale: 1. excellent; 2. very good; 3. good; 4. fair; 5. poor), (b) the number of physically unhealthy days in the past 30 days, (c) the number of mentally unhealthy days in the past 30 days, and (d) the number of days with activity limitation in the past 30 days.^13^ The CDC HRQOL-4 instrument is free for public use, not copyrighted, and does not require permission for use or licensing fees. The CDC HRQOL-4 tool was translated from English into Japanese by a Japanese epidemiologist and an occupational physician. A Japanese version of the CDC HRQOL-4 has also been developed, and several previous studies have been conducted on Japanese workers using the same.^15,16^ Previous studies have reported that the Japanese version of the HRQOL shows good concurrent validity with the Short Form-8 (SF-8 Form), a widely used indicator of HRQOL in Japan and the work functioning impairment scale (WFun), an indicator of the worker’s functional disability at work due to health problems which has good construct validity among Japanese workers.^15^ The forward translations were reconciled. After that, the back translation was performed by a native English-speaking researcher to ensure the equivalence between the original English version and the Japanese translated version. No conceptual or contextual differences from the original English version were found.^17^

The original question regarding the intensity of PWHS: “Your company supports lively working and healthy living for employees,” was asked to participants using a four-point Likert scale: strongly agree (very high PWHS), agree (high PWHS), disagree (low PWHS), and strongly disagree (very low PWHS).

### Variables

The self-rated health score, the number of physically unhealthy days in the past 30 days, the number of mentally unhealthy days in the past 30 days, and the number of days with activity limitation in the past 30 days in CDC HRQOL-4 were used as outcome variables.

According to the intensity of PWHS, we divided the participants into four groups: very high, high, low, and very low (fig 1), and these variables were used as predictor variables.

The following items, surveyed using a questionnaire, were used as confounding factors. Sex, age (20-29yr, 30-39yr, 40-49yr, 50-59yr, and ≥60 years), educational background (junior or senior high school, junior college or vocational school, university or graduate school), and presence of illnesses that require hospital treatment were personal characteristics. Employment status (regular employees, managers, executives, public service workers, temporary workers, freelancers or professionals, others), company size where participants work (≤9 employees, 10-49, 50-99, 100-499, 500-999, 1000-9999, ≥10000), working hours per day (<8h/d, 8≤ and < 9h/d, 9≤ and <11h/d, ≥11h/d) were used as work-related factors.

### Statistical Method

We used a linear mixed model (LMM) to analyze the relationship between the four groups of PWHS and the four domains of CDC HRQOL in the two models. The dependent variable from the score for each of the four domains of the CDC HRQOL-4. In the sex-age-adjusted model LMM, we added sex and age as fixed effects. In the multivariate model LMM, we added variables related to personal characteristics and work-related factors as fixed effects. Residual maximum likelihood (REML) estimation was used estimations for fixed effects in LMM. In all tests, the threshold for significance was set at p<0.05. SPSS25.0J analytical software (IBM, NY) was used.

## Results

### Participants and Descriptive Data

According to PWHS scores, there were 2,370 participants in very high PWHS, 13,159 in high PWHS, 6,941 in low PWHS, and 4,566 in very low PWHS. (Fig. 1).

Male participants had a lower proportion of very high PWHS and a higher proportion of very low PWHS comparing female participants. In participants of 20-29 years and ≥60 years, the proportion of very high PWHS was high, whereas in 40-49 years and 50-59 years, the very low PWHS was high. The proportion of participants with illnesses who required hospital treatment tended to be higher in the group with very low PWHS. For work-related factors, very high PWHS were high among participants who worked at a company size of ≥10,000 employees and those who worked < 8h/d. The proportion of very low PWHS was higher among those who worked at a company size of ≤99 employees and those who worked ≥9 h/d (Table 1).

**Table 1.**
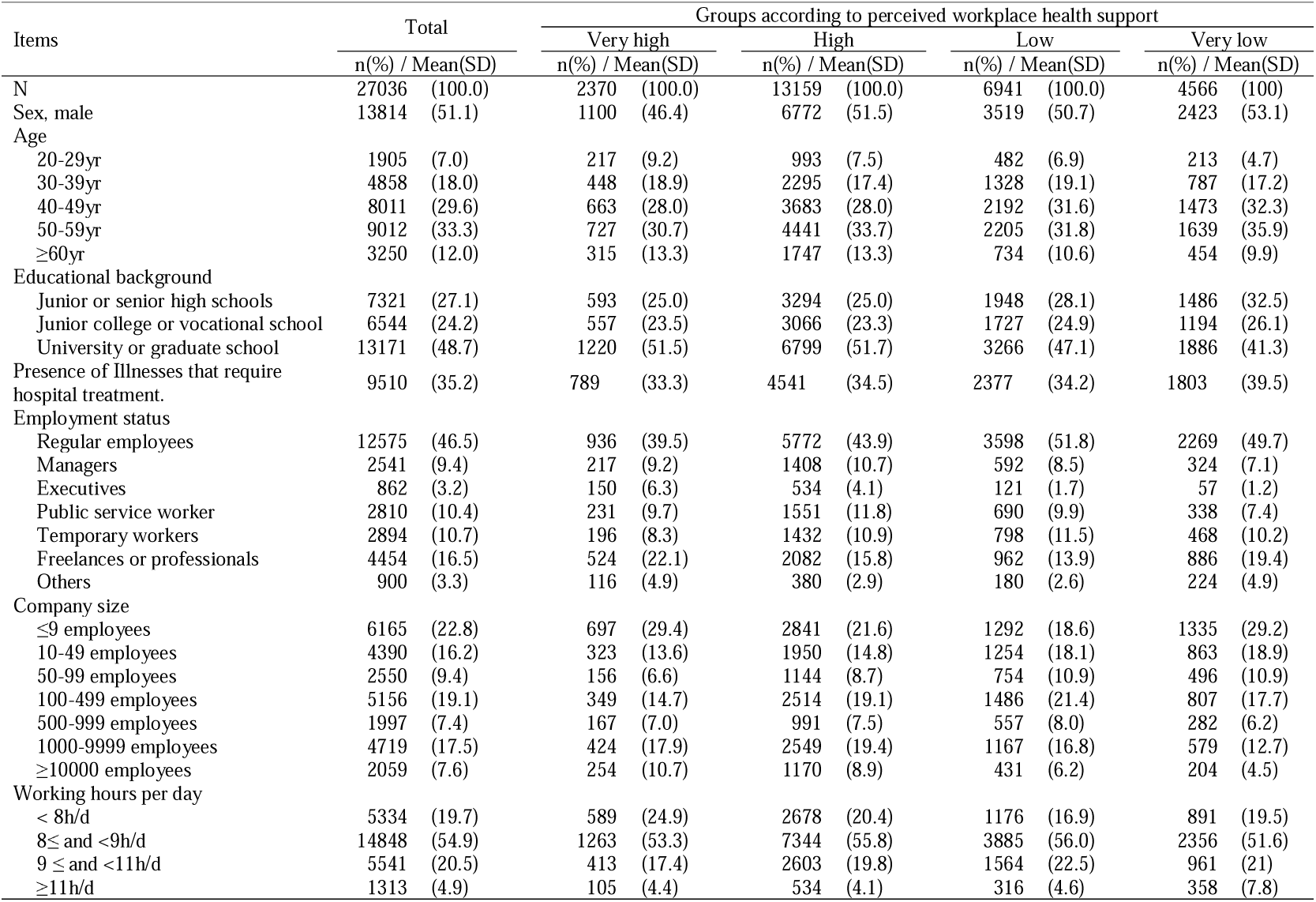
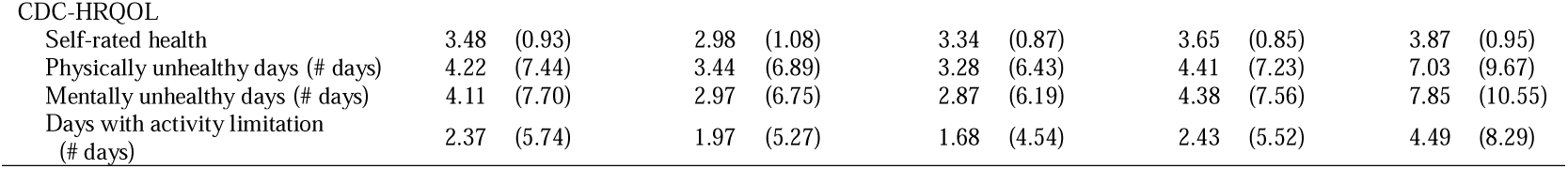
Participants’ characteristics by groups according to perceived workplace health support (PWHS)

| Items | Total |  | Groups according to perceived workplace health support |  |  |  |  |  |  |  |
| --- | --- | --- | --- | --- | --- | --- | --- | --- | --- | --- |
|  |  |  | Very high |  | High |  | Low |  | Very low |  |
|  | n(%) / Mean(SD) |  | n(%) / Mean(SD) |  | n(%) / Mean(SD) |  | n(%) / Mean(SD) |  | n(%) / Mean(SD) |  |
| N | 27036 | (100.0) | 2370 | (100.0) | 13159 | (100.0) | 6941 | (100.0) | 4566 | (100) |
| Sex, male | 13814 | (51.1) | 1100 | (46.4) | 6772 | (51.5) | 3519 | (50.7) | 2423 | (53.1) |
| Age |  |  |  |  |  |  |  |  |  |  |
| 20-29yr | 1905 | (7.0) | 217 | (9.2) | 993 | (7.5) | 482 | (6.9) | 213 | (4.7) |
| 30-39yr | 4858 | (18.0) | 448 | (18.9) | 2295 | (17.4) | 1328 | (19.1) | 787 | (17.2) |
| 40-49yr | 8011 | (29.6) | 663 | (28.0) | 3683 | (28.0) | 2192 | (31.6) | 1473 | (32.3) |
| 50-59yr | 9012 | (33.3) | 727 | (30.7) | 4441 | (33.7) | 2205 | (31.8) | 1639 | (35.9) |
| ≥60yr | 3250 | (12.0) | 315 | (13.3) | 1747 | (13.3) | 734 | (10.6) | 454 | (9.9) |
| Educational background |  |  |  |  |  |  |  |  |  |  |
| Junior or senior high schools | 7321 | (27.1) | 593 | (25.0) | 3294 | (25.0) | 1948 | (28.1) | 1486 | (32.5) |
| Junior college or vocational school | 6544 | (24.2) | 557 | (23.5) | 3066 | (23.3) | 1727 | (24.9) | 1194 | (26.1) |
| University or graduate school | 13171 | (48.7) | 1220 | (51.5) | 6799 | (51.7) | 3266 | (47.1) | 1886 | (41.3) |
| Presence of illnesses that require hospital treatment. | 9510 | (35.2) | 789 | (33.3) | 4541 | (34.5) | 2377 | (34.2) | 1803 | (39.5) |
| Employment status |  |  |  |  |  |  |  |  |  |  |
| Regular employees | 12575 | (46.5) | 936 | (39.5) | 5772 | (43.9) | 3598 | (51.8) | 2269 | (49.7) |
| Managers | 2541 | (9.4) | 217 | (9.2) | 1408 | (10.7) | 592 | (8.5) | 324 | (7.1) |
| Executives | 862 | (3.2) | 150 | (6.3) | 534 | (4.1) | 121 | (1.7) | 57 | (1.2) |
| Public service worker | 2810 | (10.4) | 231 | (9.7) | 1551 | (11.8) | 690 | (9.9) | 338 | (7.4) |
| Temporary workers | 2894 | (10.7) | 196 | (8.3) | 1432 | (10.9) | 798 | (11.5) | 468 | (10.2) |
| Freelances or professionals | 4454 | (16.5) | 524 | (22.1) | 2082 | (15.8) | 962 | (13.9) | 886 | (19.4) |
| Others | 900 | (3.3) | 116 | (4.9) | 380 | (2.9) | 180 | (2.6) | 224 | (4.9) |
| Company size |  |  |  |  |  |  |  |  |  |  |
| ≤9 employees | 6165 | (22.8) | 697 | (29.4) | 2841 | (21.6) | 1292 | (18.6) | 1335 | (29.2) |
| 10-49 employees | 4390 | (16.2) | 323 | (13.6) | 1950 | (14.8) | 1254 | (18.1) | 863 | (18.9) |
| 50-99 employees | 2550 | (9.4) | 156 | (6.6) | 1144 | (8.7) | 754 | (10.9) | 496 | (10.9) |
| 100-499 employees | 5156 | (19.1) | 349 | (14.7) | 2514 | (19.1) | 1486 | (21.4) | 807 | (17.7) |
| 500-999 employees | 1997 | (7.4) | 167 | (7.0) | 991 | (7.5) | 557 | (8.0) | 282 | (6.2) |
| 1000-9999 employees | 4719 | (17.5) | 424 | (17.9) | 2549 | (19.4) | 1167 | (16.8) | 579 | (12.7) |
| ≥10000 employees | 2059 | (7.6) | 254 | (10.7) | 1170 | (8.9) | 431 | (6.2) | 204 | (4.5) |
| Working hours per day |  |  |  |  |  |  |  |  |  |  |
| < 8h/d | 5334 | (19.7) | 589 | (24.9) | 2678 | (20.4) | 1176 | (16.9) | 891 | (19.5) |
| 8≤ and <9h/d | 14848 | (54.9) | 1263 | (53.3) | 7344 | (55.8) | 3885 | (56.0) | 2356 | (51.6) |
| 9 ≤ and <11h/d | 5541 | (20.5) | 413 | (17.4) | 2603 | (19.8) | 1564 | (22.5) | 961 | (21) |
| ≥11h/d | 1313 | (4.9) | 105 | (4.4) | 534 | (4.1) | 316 | (4.6) | 358 | (7.8) |

| Items | Groups according to perceived workplace health support |  |  |  |  |  |  |  |  |  |
| --- | --- | --- | --- | --- | --- | --- | --- | --- | --- | --- |
|  | Total |  | Very high |  | High |  | Low |  | Very low |  |
|  | n(%) / Mean(SD) |  | n(%) / Mean(SD) |  | n(%) / Mean(SD) |  | n(%) / Mean(SD) |  | n(%) / Mean(SD) |  |
| CDC-HRQOL |  |  |  |  |  |  |  |  |  |  |
| Self-rated health | 3.48 | (0.93) | 2.98 | (1.08) | 3.34 | (0.87) | 3.65 | (0.85) | 3.87 | (0.95) |
| Physically unhealthy days (# days) | 4.22 | (7.44) | 3.44 | (6.89) | 3.28 | (6.43) | 4.41 | (7.23) | 7.03 | (9.67) |
| Mentally unhealthy days (# days) | 4.11 | (7.70) | 2.97 | (6.75) | 2.87 | (6.19) | 4.38 | (7.56) | 7.85 | (10.55) |
| Days with activity limitation (# days) | 2.37 | (5.74) | 1.97 | (5.27) | 1.68 | (4.54) | 2.43 | (5.52) | 4.49 | (8.29) |

### Comparison of the Scores of the Japanese version of the CDC HRQOL-4 among Groups according to PWHS

In the mean score (SD) of self-rated health of the CDC HEQOL, the very low group was the highest of 3.87 (0.95) and the very high group had the lowest score of 2.98 (1.01). In mean (SD) of each of the three domains of: the number of physically unhealthy days, the number of mentally unhealthy days, and the number of days with activity limitation, the very low group was the highest of 7.03 (9.67), 7.85 (10.55), and 4.49 (8.29) and high group was the lowest of 3.28 (6.43), 2.87 (6.19), and 1.68 (4.54). (Table 1)

We statistically compared each score of four domain of the CDC HRQOL-4 among the PWHS groups (Table 2). In the sex-age-adjusted and multivariate-adjusted model LMM, there was a significant effect of the PWHS group on self-rated health and the self-rated health score was significantly lower (i.e. a more favorable self-rated health health) in the groups with higher PWHS.

**Table 2.** Comparison of the scores of the CDC HEQOL among groups according to perceived workplace health support (PWHS)

| Parameters | Groups according to PWHS | Sex-age adjusted |  |  | Multivariate* |  |  |
| --- | --- | --- | --- | --- | --- | --- | --- |
|  |  | Coefficients | 95%CI | p | Coefficients | 95%CI | p |
| Self-rated health | Very high | -0.89 | [-0.94– -0.85] | <0.001 | -0.85 | [-0.89– -0.81] | <0.001 |
|  | High | -0.53 | [-0.56– -0.5] | <0.001 | -0.50 | [-0.53– -0.47] | <0.001 |
|  | Low | -0.22 | [-0.25– -0.18] | <0.001 | -0.20 | [-0.23– -0.17] | <0.001 |
|  | Very low | reference |  |  | reference |  |  |
| Physically unhealthy days (# days) | Very high | -3.60 | [-3.96– -3.24] | <0.001 | -3.33 | [-3.68– -2.97] | <0.001 |
|  | High | -3.72 | [-3.97– -3.48] | <0.001 | -3.41 | [-3.65– -3.17] | <0.001 |
|  | Low | -2.64 | [-2.91– -2.36] | <0.001 | -2.35 | [-2.62– -2.09] | <0.001 |
|  | Very low | reference |  |  | reference |  |  |
| Mentally unhealthy days (# days) | Very high | -4.99 | [-5.36– -4.62] | <0.001 | -4.7 | [-5.06– -4.33] | <0.001 |
|  | High | -4.98 | [-5.23– -4.73] | <0.001 | -4.72 | [-4.97– -4.47] | <0.001 |
|  | Low | -3.54 | [-3.82– -3.26] | <0.001 | -3.33 | [-3.61– -3.06] | <0.001 |
|  | Very low | reference |  |  | reference |  |  |
| Days with activity limitation (# days) | Very high | -2.57 | [-2.85– -2.29] | <0.001 | -2.38 | [-2.66– -2.1] | <0.001 |
|  | High | -2.81 | [-3.00– -2.62] | <0.001 | -2.61 | [-2.80– -2.42] | <0.001 |
|  | Low | -2.09 | [-2.30– -1.88] | <0.001 | -1.93 | [-2.13– -1.72] | <0.001 |
|  | Very low | reference |  |  | reference |  |  |

CI: Confidence interval.
\* The multivariate model was adjusted for sex, age, educational background, presence of illnesses that require hospital treatment, employment status, company size, and working hours per day

In the sex-age-adjusted and multivariate-adjusted model LMM, there were significant consequences of the PWHS groups in each of the three domains of: the number of physically unhealthy days, the number of mentally unhealthy days, and the number of days with activity limitation (Table 2). The very low PWHS group had considerably higher values of the three domains of unhealthy days than the other three groups (p<0.001). We observed that the values of the three domains of unhealthy days of low PWHS group tended to be higher than the ones of the very high and high PWHS.

## Discussion

In this study, we analyzed the relationship between PWHS and HRQOL. We found a substantial impact of PWHS on the following four domains of CDC-HRQOL-4: self-rated health, the number of physically unhealthy days, the number of mentally unhealthy days, and the number of days with activity limitation, which tended to be worse in the group with lower PWHS. Absenteeism (the absence of a worker from work due to health problems) and presenteeism (there are several definitions, but the two main are sickness presenteeism, which is working while sick and productivity loss that stems from being at work while ill, and decreasing performance) are well-known indicators related to the health status of workers.^18,19^ Regarding the relationship between PWHS and indicators of absenteeism and presenteeism on productivity loss, Chen et al. have reported that presenteeism differs substantially with PWHS levels. People with lower PWHS had higher presenteeism than those with higher PWHS, and that higher PWHS was independently associated with higher work productivity.^6^ While, another previous study stated that PWHS, as a workplace culture with a healthy lifestyle and physical activity, was found to influence absenteeism and presenteeism through the mediation of anxiety and depressive symptoms.^20^ This study is consistent with previous reports and found that with a decrease in PWHS, self-rated health status worsens and unhealthy days increase.

In this study, the group with very low PWHS had a higher proportion of participants with illnesses who required hospital treatment and those who had working hours of ≥9 h/d than the other three groups. The companies committed to H&PM could implement workplace health promotion programs for primary prevention ^21^ or proper working hour management to prevent various diseases associated with long working hours.^22^ It has been reported that workplace elements such as environmental and policy support may be associated with a marginally significant lower lifestyle risk.^23^ We suppose that it is crucial to improve PWHS by promoting H&PM as low PWHS may hinder workers’ health.

This study was conducted in December 2020, during the COVID-19 pandemic, and there are concerns that the psychological impact of the COVID-19 pandemic may worsen the health status of individuals. This study suggests that efforts to improve PWHS in companies could help reduce the deterioration of health status even during the pandemic because of the relationship between PWHS and self-rated health or unhealthy days. In a study of occupational health, safety policies, and perceived support from the organization, supervisors, and coworkers in a petrochemical company, perceived supervisor support was reported to have a significant influence on workers’ compliance behavior.^24^ We speculate that by increasing PWHS, workers’ awareness of preventive behaviors against COVID-19 infection can be improved.

As mentioned above, there is a relationship between PWHS and the health status of workers, and it is essential to promote H&PM with higher PWHS. We believe that assessing the PWHS of workers is effective in confirming the degree of promotion of H&PM. Additionally, evaluating the use of HRQOL may be plausible as one of the outputs for health management in the workplace.

### Limitation

This study has three limitations. First, this is an internet-based survey, so the generalizability of the results is uncertain. However, to reduce sampling bias, sampling was conducted through sex, generation, personal characteristics and occupation. Second, because this was a cross-sectional study, the causal relationship between PWHS and HRQOL is unclear. However, based on previous studies that report lower PWHS affects presenteeism^6,20^, we believe that it is likely that lower PWHS leads to lower HRQOL. Third, although this study was conducted under the COVID-19 pandemic in Japan, there are differences in the participants’ awareness of PWHS and HRQOL between the present and regular times. In addition, because of the possible health concerns associated with the COVID-19 pandemic, further studies are necessary.

## Conclusion

In this study, we investigated the relationship between PWHS and HRQOL. It was found that the higher the PWHS of Japanese workers, the higher their self-rated health and the lower their unhealthy days. We believe that it is significant for companies to assess the PWHS and HRQOL of their workers to promote H&PM, as it is vital for maintaining and promoting workers’ health during the COVID-19 epidemic.

## Data Availability

Due to the nature of this research, the participants of this study did not consent to their data being publicly shared; hence, supporting data will not be made available.

## Acknowledgments

The current members of the CORoNaWork Project, in alphabetical order, are as follows: Dr. Yoshihisa Fujino (present chairperson of the study group), Dr. Akira Ogami, Dr. Arisa Harada, Dr. Ayako Hino, Dr. Hajime Ando, Dr. Hisashi Eguchi, Dr. Kazunori Ikegami, Dr. Kei Tokutsu, Dr. Keiji Muramatsu, Dr. Koji Mori, Dr. Kosuke Mafune, Dr. Kyoko Kitagawa, Dr. Masako Nagata, Dr. Mayumi Tsuji, Ms. Ning Liu, Dr. Rie Tanaka, Dr. Ryutaro Matsugaki, Dr. Seiichiro Tateishi, Dr. Shinya Matsuda, Dr. Tomohiro Ishimaru, and Dr. Tomohisa Nagata. All members are affiliated with the University of Occupational and Environmental Health, Japan.

## Funding

This study was supported and partly funded by the research grant from the University of Occupational and Environmental Health, Japan (no grant number); Japanese Ministry of Health, Labour and Welfare (H30-josei-ippan-002, H30-roudou-ippan-007, 19JA1004, 20JA1006, 210301-1, and 20HB1004); Anshin Zaidan (no grant number), the Collabo-Health Study Group (no grant number), and Hitachi Systems, Ltd. (no grant number) and scholarship donations from Chugai Pharmaceutical Co., Ltd. (no grant number)

## Conflict of interests

The authors have no conflicts of interest to declare regarding this study.

## Notes

### Competing Interest Statement

The authors have declared no competing interest.

### Author Declarations

This study was approved by the Ethics Committee of the University of Occupational and Environmental Health, Japan.

## References

1) Statistics Bureau of Japan. Statistical handbook of Japan 2019. https://www.stat.go.jp/english/data/handbook/c0117.html. Accessed April 20, 2021.

2) Ministry of Economy, Trade and Industry,. Enhancing health and productivity management. https://www.meti.go.jp/policy/mono_info_service/healthcare/downloadfiles/180717health-and-productivity-management.pdf. Accessed April 21, 2021.

3) Mori K, Nagata T, Nagata M, et al. Development, Success Factors, and Challenges of Government-Led Health and Productivity Management Initiatives in Japan. J Occup Environ Med. Jan 1 2021;63(1):18–26.

4) Eisenberger R, Huntington R, Hutchison S, Sowa D. Perceived organizational support. Journal of Applied psychology. 1986;71(3):500.

5) Allen DG, Shore LM, Griffeth RW. The role of perceived organizational support and supportive human resource practices in the turnover process. Journal of management. 2003;29(1):99–118.

6) Chen L, Hannon PA, Laing SS, et al. Perceived workplace health support is associated with employee productivity. Am J Health Promot. Jan-Feb 2015;29(3):139–146.

7) Vindegaard N, Benros ME. COVID-19 pandemic and mental health consequences: Systematic review of the current evidence. Brain, behavior, and immunity. 2020;89:531–542.

8) Giorgi G, Lecca LI, Alessio F, et al. COVID-19-related mental health effects in the workplace: a narrative review. International journal of environmental research and public health. 2020;17(21):7857.

9) Ammar A, Trabelsi K, Brach M, et al. Effects of home confinement on mental health and lifestyle behaviours during the COVID-19 outbreak: insights from the ECLB-COVID19 multicentre study. Biol Sport. Mar 2021;38(1):9–21.

10) Ammar A, Brach M, Trabelsi K, et al. Effects of COVID-19 home confinement on eating behaviour and physical activity: results of the ECLB-COVID19 international online survey. Nutrients. 2020;12(6):1583.

11) Whoqol Group. The World Health Organization quality of life assessment (WHOQOL): position paper from the World Health Organization. Social science & medicine. 1995;41(10):1403–1409.

12) Fujino Y, Ishimaru T, Eguchi H, et al. Protocol for a Nationwide Internet-based Health Survey of Workers During the COVID-19 Pandemic in 2020. Journal of UOEH. 2021;43(2):217–225.

13) Centers for Disease Control and Prevention. Measuring Healthy Days. https://www.google.com/url?sa=t&rct=j&q=&esrc=s&source=web&cd=&ved=2ahUKEwi8-ZGq8_XwAhWV_WEKHZnkBl0QFjABegQIBBAD&url=https%3A%2F%2Fwww.cdc.gov%2Fhrqol%2Fpdfs%2Fmhd.pdf&usg=AOvVaw3cH2DxYlYGGLT_3S-wc-5s. Accessed April 23, 2021.

14) Centers for Disease Control and Prevention. Measuring Healthy Days. http://www.cdc.gov/hrqol. Accessed May 19, 2021.

15) Chimed-Ochir O, Mine Y, Okawara M, Ibayashi K, Miyake F, Fujino Y. Validation of the Japanese version of the CDC HRQOL-4 in workers. Journal of Occupational Health. 2020;62(1).

16) Chimed-Ochir O, Mine Y, Fujino Y. Pain, unhealthy days and poor perceived health among Japanese workers. J Occup Health. Jan 2020;62(1):e12092.

17) Group W. The World Health Organization quality of life assessment (WHOQOL): position paper from the World Health Organization. Social science & medicine. 1995;41(10):1403–1409.

18) Gosselin E, Lemyre L, Corneil W. Presenteeism and absenteeism: differentiated understanding of related phenomena. J Occup Health Psychol. Jan 2013;18(1):75–86.

19) Johns G. Presenteeism in the workplace: A review and research agenda. Journal of organizational behavior. 2010;31(4):519–542.

20) Laing SS, Jones SM. Anxiety and Depression Mediate the Relationship Between Perceived Workplace Health Support and Presenteeism: A Cross-sectional Analysis. J Occup Environ Med. Nov 2016;58(11):1144–1149.

21) Goetzel RZ, Ozminkowski RJ. Health and productivity management: emerging opportunities for health promotion professionals for the 21st century. American Journal of Health Promotion. 2000;14(4):211–214.

22) Bannai A, Tamakoshi A. The association between long working hours and health: a systematic review of epidemiological evidence. Scand J Work Environ Health. Jan 2014;40(1):5–18.

23) Payne J, Cluff L, Lang J, Matson-Koffman D, Morgan-Lopez A. Elements of a Workplace Culture of Health, Perceived Organizational Support for Health, and Lifestyle Risk. Am J Health Promot. Sep 2018;32(7):1555–1567.

24) Puah LN, Ong LD, Chong WY. The effects of perceived organizational support, perceived supervisor support and perceived co-worker support on safety and health compliance. Int J Occup Saf Ergon. Sep 2016;22(3):333–339.

